# A clinical algorithm to identify people with the glucose-6-phosphate dehydrogenase p.Val68Met variant at risk for diabetes undertreatment

**DOI:** 10.1101/2025.01.17.25320736

**Authors:** Yash Pershad, Joseph H Breeyear, Robert W Corty, Jonathan D Mosley, Ayush Giri, Dan M Roden, Todd L Edwards, Alexander G Bick

## Abstract

**Purpose:** To develop an algorithm using routine clinical laboratory measurements to identify people at risk for systematic underestimation of glycated hemoglobin (HbA1c) due to p.Val68Met glucose-6-phosphate dehydrogenase (G6PD) deficiency.

**Methods:** We analyzed 122,307 participants of self-identified Black race across four large cohorts with blood glucose, HbA1c, and red cell distribution width measurements from a single blood draw. In UK Biobank, we used recursive partitioning to develop criteria for possible and likely G6PD deficiency. We validated the algorithm in NIH All of Us, Vanderbilt BioVU, and the Million Veterans Program. In Vanderbilt’s Synthetic Derivative, we created a cohort of 48,031 participants with type 2 diabetes and no genetic data to test whether predicted risk for G6PD deficiency was associated with incident diabetic retinopathy.

**Results:** G6PD deficiency predictions in hemizygous males showed precision/recall of 31%/81% for possible and 81%/10% for likely deficiency. In homozygous females, precision/recall was 6%/76% for possible and 34%/13% for likely deficiency. Diabetic patients with predicted possible deficiency demonstrated 1.4-fold higher 20-year retinopathy rates (14.3% vs 11.2%, P=0.003).

**Conclusion:** We report a simple clinical algorithm that enables healthcare systems to identify people who may benefit from G6PD genotyping and glucose-based diabetes monitoring.

## Introduction

The promise of translating genetic insights into clinical medicine remains partially unfulfilled, particularly for historically underserved populations. The goal of a genomic learning health system (gLHS) is to identify and advance approaches for integrating genomic information into existing learning health systems. A major barrier to achieving this goal is the present inability to perform genetic testing in all people. Therefore, a gLHS requires tools to quantify the burden of undiagnosed genetic conditions within its patient populations and prioritize those who may benefit from genetic testing.

An example of this situation is glucose-6-phosphate dehydrogenase (G6PD) deficiency. This X-linked enzymopathy, which shortens erythrocyte lifespan,^1,2^ causes systematically lower glycated hemoglobin (HbA1c) relative to blood glucose,^3,4^ creating a population-level risk of diabetes underdiagnosis and undertreatment that disproportionately affects people of African ancestry.^5^ The American Diabetes Association standard of care in diabetes identifies G6PD deficiency as a major cause of interference with HbA1c reference ranges and recommends to use plasma glucose to diagnose and manage diabetes. ^6^ Despite the prevalence of G6PD deficiency and the knowledge of its impact on diabetes management, people with G6PD deficiency typically remain unidentified unless they experience rare hemolytic crises.^2,7^ Since we are currently unable to test all people for G6PD deficiency, there is a critical need for a tool to identifying high-risk people who should be tested.

Here, we developed an algorithm using routine laboratory measurements to estimate G6PD p.Val68Met variant prevalence and prioritize a group who would likely benefit from genotyping for the variant. The G6PD p.Val68Met variant, found almost exclusively in those of African ancestry, demonstrates how algorithms built with clinical data can help gLHSs identify vulnerable populations to prioritize initial genetic-based interventions.

## Materials and Methods

We analyzed data from 122,307 participants of self-identified Black race with concurrent blood glucose, HbA1c, and red cell distribution width (RDW) measurements across three cohorts: UK Biobank (N=3,864), NIH All of Us (N=2,047), Vanderbilt BioVU (N=2,087), and the Million Veterans Program (MVP) (N=114,309). We limited our analysis within the cohorts to those of self-identified Black race because clinicians applying this algorithm will be presently unaware of genetic ancestry. Furthermore, the G6PD p.Val68Met variant is almost exclusively found in those of self-identified Black race. Zygosity of the G6PD p.Val68Met variant was determined by extracting genotype for variant rs1050828-T.

In the UK Biobank, blood glucose (Data-Field 30740), HbA1c (Data-Field 30750), and RDW (Data-Field 30070) were measured for all participants at baseline with the same blood draw. UK Biobank participants were asked to fast before blood glucose measurements. In All of Us, BioVU, and MVP, blood glucose (OMOP ID: 3004501), HbA1c (OMOP ID: 3004410), and RDW (OMOP ID: 3019897) were obtained as part of clinical care and linked to participant data. Since these measurements were not assured to be fasting, blood draws with blood glucose measurements less than 70 mg/dL and greater than 200 mg/dL were excluded. If an All of Us, BioVU, or MVP participant had more than one eligible blood draw, the HbA1c, RDW, and blood glucose values from the draw with the lowest blood glucose (most likely to be fasting) was used. We calculated the glucose gap (GG) as measured blood glucose minus the estimated average glucose level as calculated from HbA1c (28.7 x HbA1c - 46.7).8 Clinical diagnosis of G6PD deficiency and diabetes mellitus were defined by ICD codes (**Supplementary Table 1**).

We evaluated the algorithm’s ability to discern three patient populations: 1) male G6PD p.Val68Met hemizygotes, 2) female G6PD p.Val68Met homozygotes, and 3) females with at least one G6PD p.Val68Met allele (heterozygotes and homozygotes). Using recursive partitioning in the UK Biobank (scikit-learn v1.5.1), we developed criteria for “likely” G6PD deficiency (both RDW ≤12 fL and GG ≥-15 mg/dL) and “possible” G6PD deficiency (either RDW ≤12 fL or GG ≥-15 mg/dL). We assessed precision 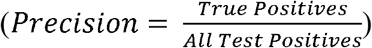 and recall 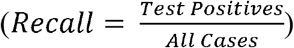 of possible and likely G6PD deficiency criteria for each of the three tasks in all cohorts.^9^ For precision, true positives were defined as participants meeting the likely or possible criteria with the relevant genotype, while test positives were all participants meeting the likely or possible criteria regardless of genotype. For recall, true positives were defined as above, while all cases were all participants with the relevant genotype.

We then demonstrated population-level application of the algorithm by analyzing 48,031 people of self-identified Black race with type 2 diabetes in Vanderbilt’s Synthetic Derivative who did not have genetic data available. This health system implementation allowed us to estimate the burden of potentially undiagnosed G6PD deficiency and identify subpopulations at elevated risk for diabetes complications. We calculated population-level glucose gaps and applied our thresholds to estimate G6PD p.Val68Met status. Using Cox proportional hazards regression, we examined time (years) from diabetes diagnosis to incident diabetic retinopathy diagnosis, stratified by predicted G6PD deficiency burden. Follow-up was censored at time of diabetic retinopathy diagnosis (ICD codes in **Supplementary Table 1**), death, or end of follow-up, whichever came first.

## Results

Among UK Biobank participants who self-identify as Black race, 15.4% of males were p.Val68Met hemizygotes, while 26.4% and 2.4% of females were heterozygotes and homozygotes, respectively. Similar frequencies were observed in All of Us (8.8% hemizygotes, 20.6% heterozygotes, 0.8% homozygotes), BioVU (10.3% hemizygotes, 20.1% heterozygotes, 1.8% homozygotes), and MVP (11.7% hemizygotes, 21.0% heterozygotes, and 1.4% homozygotes). Prevalence of G6PD p.Val68Met alleles was highest in the UK Biobank. Notably, <0.5% of G6PD p.Val68Met hemizygotes and homozygotes across all cohorts had a clinical G6PD deficiency diagnosis (**Table 1**).

**Table 1.** Laboratory characteristics, G6PD p.Val68Met variant frequencies, and clinical diagnoses across four cohorts, stratified by sex. Values are presented as median ± interquartile range (IQR) for laboratory measurements and N (%) for genotypes and diagnoses. The glucose gap represents the difference between measured blood glucose and HbA1c-estimated glucose. N/A indicates not applicable for male heterozygote status since *G6PD* is on chromosome X and the clinical diagnoses of G6PD deficiency in MVP which were not available.

|  | UK Biobank<br>(N = 3,864) |  | All of Us<br>(N = 2,047) |  | BioVU<br>(N=2,087) |  | MVP<br>(N=114,309) |  |
| --- | --- | --- | --- | --- | --- | --- | --- | --- |
|  | Males<br>(N=1,914) | Females<br>(N=1,950) | Males<br>(N=568) | Females<br>(N=1,479) | Males<br>(N=856) | Females<br>(N=1,231) | Males<br>(N=99,312) | Females<br>(N=14,997) |
| **Labs** |  |  |  |  |  |  |  |  |
| Hemoglobin A1c<br>(%, median ± IQR) | 5.6 ± 0.6 | 5.6 ± 0.6 | 5.9 ± 1.0 | 5.8 ± 0.9 | 5.9 ± 1.3 | 6.0 ± 1.4 | 6.0 ± 1.1 | 5.5 ± 0.8 |
| Blood glucose<br>(mg/dL, median ± IQR) | 88.0 ± 14.7 | 86.7 ± 13.1 | 102.0 ± 27.0 | 97.0 ± 24.0 | 101.0 ± 28.0 | 101.0 ± 31.0 | 105.3 ± 26.7 | 96.8 ± 18.7 |
| Glucose gap (mg/dL, median ± IQR) | -26.0 ± 20.1 | -25.8 ± 19.5 | -20.8 ± 26.9 | -21.0 ± 22.8 | -19.9 ± 31.8 | -23.8 ± 27.3 | -21.2 ± 20.6 | -22.5 ± 16.6 |
| Red cell distribution width<br>(fL, median ± IQR) | 13.8 ± 1.2 | 14.0 ± 1.5 | 13.8 ± 1.5 | 14.1 ± 1.9 | 14.0 ± 1.7 | 14.3 ± 2.0 | 14.0 ± 1.6 | 14.0 ± 1.5 |
| **G6PD p.Val68Met genotype** |  |  |  |  |  |  |  |  |
| Heterozygotes – (%) | N/A | 514 (26.4%) | N/A | 304 (20.6%) | N/A | 248 (20.1%) | N/A | 3,142 (21.0%) |
| Hemi/homozygotes – (%) | 294 (15.4%) | 46 (2.4%) | 50 (8.8%) | 12 (0.8%) | 88 (10.3%) | 22 (1.8%) | 11,610 (11.7%) | 208 (1.4%) |
| **Clinical Diagnoses** |  |  |  |  |  |  |  |  |
| G6PD Deficiency – (%) | 0 (0.0%) | 0 (0.0%) | 0 (0.0%) | 5 (0.3%) | 3 (0.4%) | 1 (0.1%) | N/A | N/A |
| Diabetes Mellitus – (%) | 106 (5.5%) | 72 (3.7%) | 314 (55.3%) | 738 (49.9%) | 484 (56.5%) | 771 (62.6%) | 32,802 (33.0%) | 3,391 (22.6%) |

In the UK Biobank, RDW and GG showed strong discriminative ability for male hemizygotes (AUROC: 0.82 ± 0.03) and female homozygotes (AUROC: 0.78 ± 0.04), with moderate performance for either female heterozygotes or homozygotes (AUROC: 0.63 ± 0.02). Using recursive partitioning in the UK Biobank, we identified optimal thresholds of RDW ≤12 fL and GG ≥-15 mg/dL for detecting G6PD deficiency.

We defined two classification thresholds: “possible” G6PD deficiency (meeting either criterion) or “likely” G6PD deficiency (meeting both criteria). For male hemizygotes, possible G6PD deficiency showed high recall (81.0%) with moderate precision (31.1%), while likely G6PD deficiency offered higher precision (80.8%) but lower recall (9.9%). Among females, possible G6PD deficiency identified heterozygotes or homozygotes with 50.3% recall and 43.3% precision, whereas likely G6PD deficiency showed 3.1% recall and 65.6% precision. For female homozygotes, recall and precision for possible G6PD deficiency were 76.2% and 6.2% and for likely G6PD deficiency were 13.4% and 33.9% (**Table 2**).

**Table 2.**
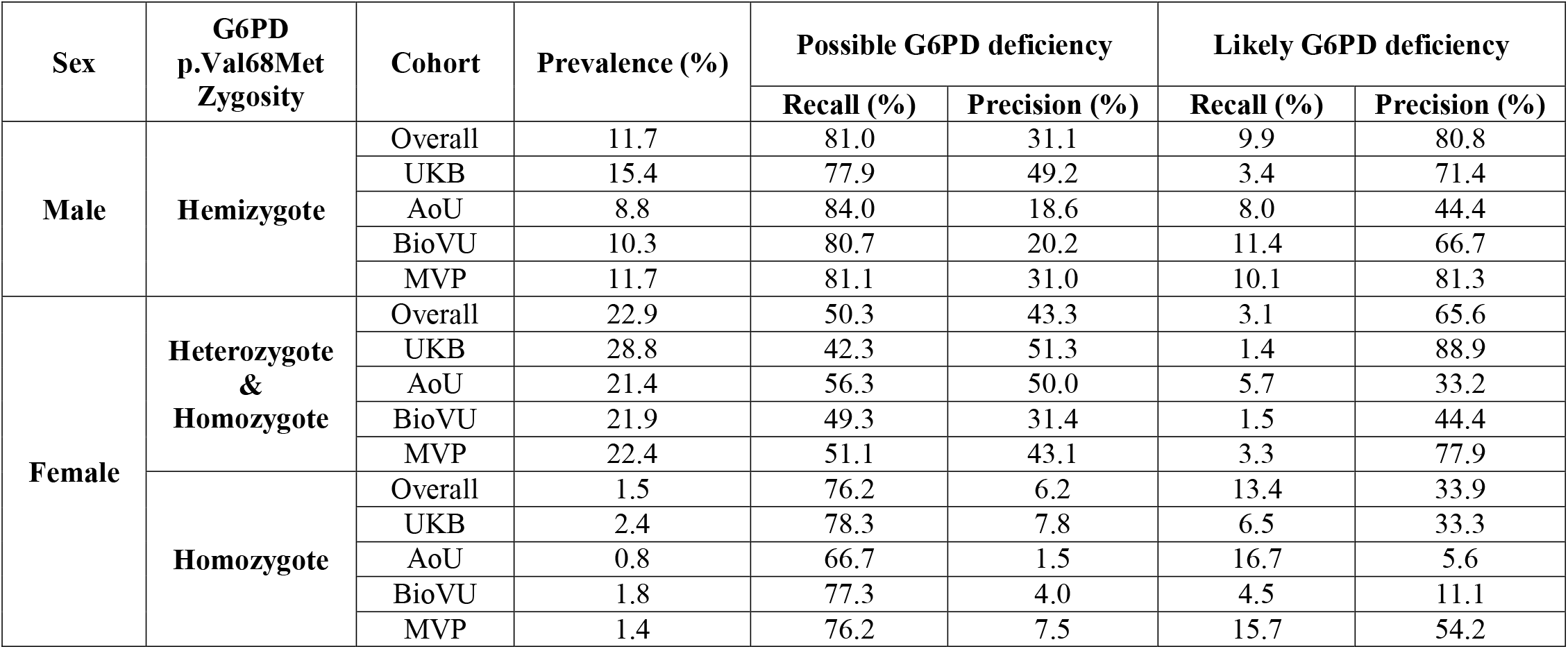
Precision and recall by sex, zygosity, and cohort for (1) possible G6PD deficiency (G6PDdef) defined as red cell distribution width ≤ 12 fL or glucose gap ≥ -15 mg/dL, and (2) likely G6PD deficiency (G6PDdef) defined as red cell distribution width ≤ 12 fL and glucose gap ≥ -15 mg/dL. UK Biobank = UKB. MVP = Million Veterans Program. All of Us = AoU.

| Sex | G6PD<br>p.Val68Met<br>Zygosity | Cohort | Prevalence (%) | Possible G6PD deficiency |  | Likely G6PD deficiency |  |
| --- | --- | --- | --- | --- | --- | --- | --- |
|  |  |  |  | Recall (%) | Precision (%) | Recall (%) | Precision (%) |
| Male | Hemizygote | Overall | 11.7 | 81.0 | 31.1 | 9.9 | 80.8 |
|  |  | UKB | 15.4 | 77.9 | 49.2 | 3.4 | 71.4 |
|  |  | AoU | 8.8 | 84.0 | 18.6 | 8.0 | 44.4 |
|  |  | BioVU | 10.3 | 80.7 | 20.2 | 11.4 | 66.7 |
|  |  | MVP | 11.7 | 81.1 | 31.0 | 10.1 | 81.3 |
| Female | Heterozygote<br>&<br>Homozygote | Overall | 22.9 | 50.3 | 43.3 | 3.1 | 65.6 |
|  |  | UKB | 28.8 | 42.3 | 51.3 | 1.4 | 88.9 |
|  |  | AoU | 21.4 | 56.3 | 50.0 | 5.7 | 33.2 |
|  |  | BioVU | 21.9 | 49.3 | 31.4 | 1.5 | 44.4 |
|  |  | MVP | 22.4 | 51.1 | 43.1 | 3.3 | 77.9 |
|  | Homozygote | Overall | 1.5 | 76.2 | 6.2 | 13.4 | 33.9 |
|  |  | UKB | 2.4 | 78.3 | 7.8 | 6.5 | 33.3 |
|  |  | AoU | 0.8 | 66.7 | 1.5 | 16.7 | 5.6 |
|  |  | BioVU | 1.8 | 77.3 | 4.0 | 4.5 | 11.1 |
|  |  | MVP | 1.4 | 76.2 | 7.5 | 15.7 | 54.2 |

To assess utility when applied at population-scale, we applied our algorithm to 48,031 people who self-identify as Black race with type 2 diabetes within our health system. The model identified substantial subpopulations requiring enhanced screening: 64.2% met possible and 1.1% met likely G6PD-deficient criteria. These groups showed higher 20-year diabetic retinopathy rates (possible: 13.9%; likely: 14.3%) compared to the general population (11.2%). After adjustment, both classifications predicted increased retinopathy risk (likely hazard ratio (HR): 1.41, 95% CI: 1.12-1.78, P=0.003; possible HR: 1.35, 95% CI: 1.28-1.42, P = 2×10-16). Both diagnoses and treatment occurred later among high-risk subgroups.

## Discussion

Recent research studies have shown those harboring the G6PD p.Val68Met variant face potential diabetes undertreatment due to their lower HbA1c levels relative to blood glucose.3,4 However, translating this finding into clinical care remains challenging because few with G6PD deficiency – we estimate < 0.5% – are aware of their enzymopathy. This proof-of-concept study demonstrates how readily available clinical data can identify vulnerable populations who may benefit from targeted screening and intervention programs. Our population health strategy offers distinct advantages for health system implementation, requiring only routine laboratory measurements while providing flexible classification thresholds for different screening scenarios. The “likely G6PD deficiency” criteria identify populations with six-fold enrichment of those with the G6PD p.Val68Met variant for focused genetic testing programs, while the “possible G6PD deficiency” criteria enable broad population-level risk assessment. Most importantly, we identified a population within the healthcare system who are currently being undertreated for diabetes – this undertreatment may be remedied by genotyping high-risk groups and using plasma glucose for diabetes monitoring, as recommended by the American Diabetes Association.^6^

While key limitations of our algorithm include reduced sensitivity for female heterozygotes due to X-chromosome inactivation,^10^ focus on a single variant in people self-identifying as Black race, our algorithm provides a model for identifying vulnerable population who may benefit from targeting genotyping. Future studies should evaluate the effectiveness and implementation outcomes of this population health approach within gLHSs. Identifying, implementing, and critically evaluating such algorithms and interventions across diverse practice settings is a crucial step to advance both genetic medicine and health equity.

## Data Availability

All data are available on the UK Biobank Access Management System https://www.ukbiobank.ac.uk/enable-your-research/apply-for-access, All of Us Researcher Workbench https://www.researchallofus.org/, Vanderbilt BioVU Terra.bio cloud environment, and Million Veterans Program Research Hub https://www.mvp.va.gov/pwa/discover-mvp-data

## Acknowledgements

We gratefully acknowledge the participants of UK Biobank, NIH All of Us, Vanderbilt BioVU, and the Million Veterans Program.

## Author Contributions

Conceptualization: Y.P., T.L.E., A.G.B.; Data curation: Y.P., J.H.B.; Formal analysis: Y.P., J.H.B.; Funding acquisition: A.G.B., T.L.E., J.D.M.; Investigation: Y.P., J.H.B., T.L.E., A.G.B., R.W.C., J.D.M.; Methodology:Y.P., J.H.B., T.L.E., A.G.B.; Project administration: A.G.B.; Resources: A.G.B., T.L.E., A.G.; Software: Y.P.,J.H.B.; Supervision: A.G.B., T.L.E.; Validation: Y.P., J.H.B., R.W.C.; Visualization: Y.P.; Writing-original draft: Y.P., A.G.B.; Writing-review & editing: All authors.

## Ethics Declaration

Individual level data was deidentified in all cohorts.

